# Testing the sensitivity of diagnosis-derived patterns in functional brain networks to symptom burden in a Norwegian youth sample

**DOI:** 10.1101/2023.10.09.23296736

**Authors:** Irene Voldsbekk, Rikka Kjelkenes, Erik R. Frogner, Lars T. Westlye, Dag Alnæs

## Abstract

Aberrant brain network development represents a putative aetiological component in mental disorders, which typically emerge during childhood and adolescence. Previous studies have identified resting-state functional connectivity (RSFC) patterns reflecting psychopathology, but the generalisability to other samples and politico-cultural contexts has not been established.

We investigated whether a previously identified cross-diagnostic case-control and autism spectrum disorder (ASD)-specific pattern of RSFC (discovery sample; aged 5-21 from New York City, USA; n=1666) could be validated in a Norwegian convenience-based youth sample (validation sample; aged 9-25 from Oslo, Norway; n=531). As a test of generalisability, we investigated if these diagnosis-derived RSFC patterns were sensitive to levels of symptom burden in both samples, based on an independent measure of symptom burden.

Both the cross-diagnostic and ASD-specific RSFC pattern were validated across samples. Connectivity patterns were significantly associated with thematically appropriate symptom dimensions in the discovery sample. In the validation sample, the ASD-specific RSFC pattern showed a weak, inverse relationship with symptoms of conduct problems, hyperactivity, and prosociality, while the cross-diagnostic pattern was not significantly linked to symptoms.

Diagnosis-derived connectivity patterns in a developmental clinical US sample were validated in a convenience sample of Norwegian youth, however, they were not associated with mental health symptoms.

## 1 Introduction

Childhood and adolescence constitute periods of life characterised by substantial developmental adaptations. These include rapid physical, hormonal, brain, cognitive, and psychological changes, adapted to the increasing complexity of our social environment and expectations with age. For example, during this time, the functional networks of the brain undergo large-scale reorganisation and maturation (Paus et al., 2008; Power et al., 2010; Sydnor et al., 2021). Adolescence is also a period with a marked increase in the incidence of psychopathology (Kessler et al., 2007). The co-occurrence of these phenomena has led to the hypothesis that increased brain plasticity during this period results in increased susceptibility to mental illness (Paus et al., 2008). Several studies have identified plausible links between psychopathology in youth and resting-state functional connectivity (RSFC) derived from functional magnetic resonance imaging (fMRI). However, the generalisability of such network patterns to vulnerability for mental illness in non-clinical samples is currently not well demonstrated.

In the context of generalisability and vulnerability, a related question is whether RSFC patterns are specific to diagnostic categories of mental disorders or shared across disorders. Considerable effort has been made to characterise RSFC patterns associated with both diagnostic syndromes and dimensional symptom scores. Transdiagnostic patterns can be identified by including participants with a range of (comorbid) diagnoses, or by modelling dimensional scores of multiple symptom domains. Using these approaches, an increasing number of studies have reported that RSFC patterns relating to psychopathology are transdiagnostic or shared across disorders (Elliott et al., 2018; Karcher et al., 2021; Kebets et al., 2023; Lees et al., 2021; Linke et al., 2021; McTeague et al., 2017; McTeague et al., 2020; Sha et al., 2019; Voldsbekk et al., 2023; Xia et al., 2018). For example, in a population-based sample of children (Adolescent Brain Cognitive Development cohort; ABCD), a shared psychopathology factor was derived and linked to RSFC using both symptom data (Karcher et al., 2021) and diagnostic data (Lees et al., 2021).

The convergence of studies on a shared factor across disorders from studies using both symptom scores as well as binary diagnosis information supports the notion of a latent vulnerability factor on which the diagnostic categories represent extremes (Sprooten et al., 2022). The above-mentioned findings linking a shared latent mental illness factor to brain measures are promising with regard to detecting neural signatures of psychopathology risk in the youth brain. However, an important question is whether these clinical RSFC patterns are sensitive to symptom burden and by extension putative risk in youth samples that are not enriched with mental disorder diagnoses.

Recently, we estimated both diagnosis-specific and cross-diagnostic RSFC patterns in a clinical developmental sample. Investigating shared associations across RSFC data and diagnostic information, we identified a pattern specific to a diagnosis of autism spectrum disorder (ASD). The other diagnosis categories did not exhibit a significant specific RSFC pattern, instead they exhibited a shared patterns across attention-deficit hyperactivity disorder (ADHD), ASD, other neurodevelopmental disorders, anxiety, mood-disorders, and other diagnoses versus no diagnosis (Voldsbekk et al., 2023). A possible interpretation of this finding is that cross-diagnostic and ASD-specific patterns represent the two most reliable RSFC markers of psychopathology. For any such patterns to be clinically relevant, they would need to show generalisability to vulnerability for mental illness, expressed as elevated clinical symptom scores, in non-clinical samples. Thus, in the current study we aimed to investigate whether the two identified diagnosis-derived RSFC patterns are sensitive to mental health symptoms in a Norwegian convenience-based sample of youth. To do this, we investigated whether a) the cross-diagnostic and ASD-specific RSFC patterns previously identified could be validated in the validation sample, and b) if these RSFC patterns associate with levels of symptom burden in the validation sample. As a further test of external validity, we also tested whether c) the RSFC patterns previously identified associate with levels of symptom burden in the discovery sample.

## 2 Material and Methods

### 2.1 Samples

#### 2.1.1 Discovery sample - HBN

Children and adolescents from New York City, USA were recruited to be part of the HBN cohort (Alexander et al., 2017). The majority have a least one diagnosed mental disorder. In the previous study (Voldsbekk et al., 2023), 1880 participants in HBN took part. 1689 of these were in the discovery sample. Of these, 1666 had available symptom score data used for the current investigation. Missing values in the symptom data were imputed with knnimpute in MATLAB (MathWorks, 2020). To check that imputation did not influence the results, we reran the analysis in the 1610 participants without missing data and no imputation. This analysis revealed similar associations between RSFC and symptom level as the original analysis (correlations were r=.99 between imputed and non-imputed result for both the cross-diagnostic and ASD patterns). For more details regarding MRI data cleaning and quality assurance steps, see Voldsbekk et al. (2023). The final sample consisted of 1666 participants (641 females, mean ± sd age: 10.91 ± 3.14, range: 5-21). See Figure 1 and table S1 for distributions of sample characteristics.

**Figure 1.**
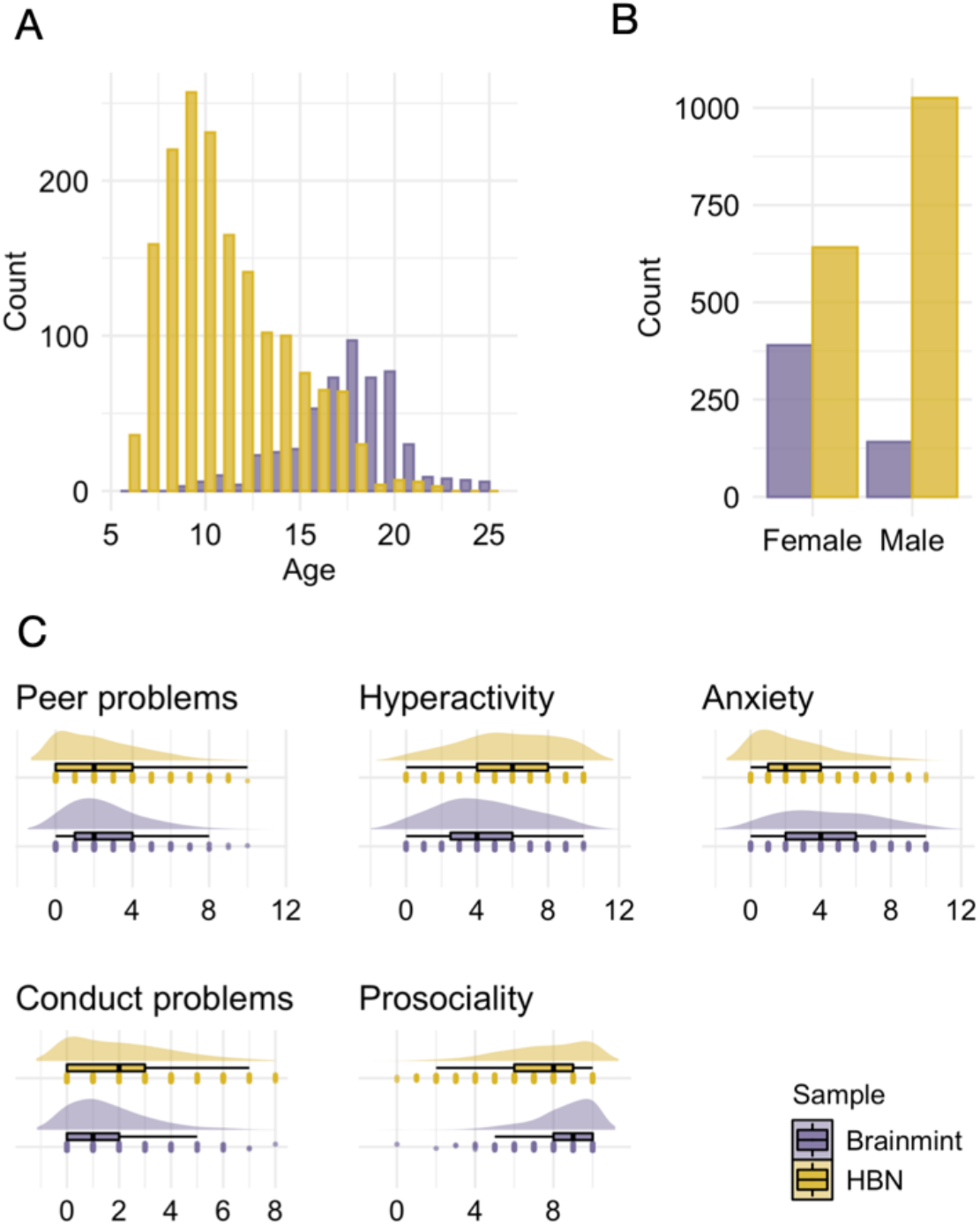
Sample distributions. **A**. Age. **B**. Sex. **C**. SDQ summary syndrome scores. HBN; Healthy brain network sample. Brainmint; Brains and minds in transition sample. SDQ; Strengths and difficulties questionnaire.

#### 2.1.2 Validation sample – Brainmint

Children and adolescents in the Oslo region were recruited to participate in the Brains and minds in transition (Brainmint) study. Participants were recruited through convenience sampling by advertising in social media, aiming to recruit young people from the general population interested in contributing to a study investigating brain development and mental health. All participants provided written informed consent prior to their participation in the study. For participants under the age of 16, both parents/legal guardians provided written informed consent on their behalf. Per May 4^th^ 2023, 759 participants had undergone magnetic resonance imaging (MRI) and 697 had responded to questionnaires. Of these, 531 had available both fMRI and symptom score data. No participants had missing data and so all were included in the sample used for the current analysis (390 females, mean ± sd age: 17.69 ± 2.83, range: 9-25). See Figure 1 and table S1 for distributions of sample characteristics.

#### 2.1.3 Mental health measures

In the previous study (Voldsbekk et al., 2023), we investigated diagnosis-derived patterns of RSFC in HBN (discovery sample). Diagnostic information was collected using a computerised version of the Schedule for Affective Disorders and Schizophrenia – Children’s version (KSADS) (Kaufman et al., 1997), which is a clinician-administered semi-structured psychiatric interview based on DSM-5. We then labelled diagnoses as belonging to either of these categories: “ADHD”, “ASD”, “anxiety disorders”, “mood disorders”, “other neurodevelopmental disorders”, “other disorders” or “no diagnosis”.

For testing the external validity and generalisability of diagnosis-derived RSFC patterns, the current study obtained HBN symptom scores from the Strength and Difficulties Questionnaire (Goodman, 1997), which is a 25-item questionnaire measuring emotional and behavioural problems. The SDQ has five syndrome measures: emotional symptoms, conduct problems, hyperactivity/inattention, peer relationship problems, and prosocial behaviour. For this analysis, we used only the summary syndrome measures. In HBN, the SDQ items were parent-reported. See Figure S1 for symptom load distributions on SDQ summary measures across diagnostic categories in HBN.

Symptom scores in Brainmint were investigated using the same approach as in HBN, only in this sample the SDQ responses were self-reported.

### 2.3 MRI acquisition

HBN MRI data were acquired at four different sites: a mobile scanner at Staten Island (SI), Rutgers University Brain Imaging Centre, Citigroup Biomedical Imaging Centre (CBIC) and Harlem CUNY Advanced Science Research Centre. A detailed overview of the MRI protocol is available elsewhere (http://fcon_1000.projects.nitrc.org/indi/cmi_healthy_brain_network/MRI%20Protocol.html).

Brainmint MRI data were acquired at Oslo University Hospital Ullevål, using a 3.0 T GE SIGNA Premier scanner using a 48-channel head coil. Structural MRI data was acquired using an T_1_-weighted MPRAGE sequence (repetition time (TR): 2.526 s, echo time (TE): 2.836 ms, flip angle (FA): 8°, field of view (FOV): 256 mm), slice thickness: 1 mm, locations per slab: 196 (no overlaps). Resting-state functional MRI (rs-fMRI) data was acquired using a T2*-weighted blood-oxygen-level-dependent echo-planar imaging (EPI) sequence with a TR of 800 ms, TE of 30 ms, multiband acceleration factor = 6, number of slices: 60, 750 repetitions and voxel size = 2.4 × 2.4 × 2.4 mm.

### 2.4 MRI pre-processing

Rs-fMRI images in HBN were processed with the following pipeline. First, FSL MCFLIRT (Jenkinson et al., 2002) was applied for motion correction, as well as high-pass temporal filtering (cut-off: 100s), spatial smoothing (FWHM: 6mm) and distortion correction as part of FEAT (Woolrich et al., 2001). The rs-fMRI images were registered to a T1-weighted structural image using FLIRT (Jenkinson et al., 2002) and boundary-based registration (Greve & Fischl, 2009). Next, for additional removal of artefacts and noise, we performed non-aggressive ICA-AROMA (Pruim, Mennes, Buitelaar, et al., 2015; Pruim, Mennes, van Rooij, et al., 2015) and FIX (Griffanti et al., 2014; Salimi-Khorshidi et al., 2014). Estimations of temporal signal-to-noise ratio (tSNR) and mean framewise displacement (FD) were calculated by MRIQC (Esteban et al., 2017) and used as covariates in subsequent analyses.

In Brainmint, preprocessing of rs-fMRI images were run using fMRIPrep v22.0.1 (Esteban et al., 2019), an automated pipeline consisting of head motion correction, high-pass temporal filtering, spatial smoothing and distortion correction using MCFLIRT, slice-timing correction using 3dTshift from AFNI, registration to structural reference image using FLIRT and boundary-based registration, and, finally, non-aggressive ICA-AROMA. Same as for HBN, estimations of tSNR and FD were calculated by MRIQC (Esteban et al., 2017).

### 2.5 Network analysis

RSFC in HBN were derived using the Schaefer parcellation with 100 parcels and 7 networks (Schaefer et al., 2018). These networks include visual A, visual B, visual C, auditory, somatomotor A, somatomotor B, language, salience A, salience B, control A, control B, control C, default A, default B, default C, dorsal attention A and dorsal attention B. The connectivity matrix was then estimated as the L2-norm ridge regression partial correlation between parcel timeseries using FSLNets (https://fsl.fmrib.ox.ac.uk/fsl/fslwiki/FSLNets), as implemented in MATLAB (MathWorks, 2020). This resulted in 4950 unique partial correlations (i.e., edges).

RSFC in Brainmint were derived using the same approach as in HBN, making the edges comparable.

### 2.6 Out-of-sample validation

In the previous work (Voldsbekk et al., 2023), we investigated diagnosis-specific RSFC patterns by running non-rotated behavioural partial least squares (PLS) in PLS Application (Krishnan et al., 2011). Non-rotated behavioural PLS yields maximal covariance across two matrices without rotating the behavioural matrix – in this case, maximal RSFC covariance across each diagnosis (a behavioural matrix containing either 1 or 0 for each participant for each diagnosis category). This test was run for each diagnosis category separately, while controlling for all other diagnosis categories using contrasts. This analysis revealed an ASD-specific pattern, as well as a cross diagnostic case-control pattern. The other diagnosis categories did not exhibit a diagnosis-specific RSFC pattern.

#### 2.6.1 Out-of-sample validation

See Figure 2 for an overview of the out-of-sample validation pipeline. First, we decomposed the Brainmint RSFC data by multiplying them with the brain weights estimated in the HBN PLS analysis. These Brainmint brain weights were then correlated with the original Brainmint RSFC data to get Brainmint connectivity loadings. Then, to assess the validation of the brain pattern across the two samples, we correlated the Brainmint connectivity loadings with HBN connectivity loadings for each latent variable (LV) using Pearson’s correlation. Their significance was tested using permutations (n=1000), randomly shuffling the rows (participants) of the Brainmint RSFC data. We calculated *p*-values by dividing the count of permuted maximum R values (including the observed non-permuted value) ≥ the non-permuted R values by the number of permutations. Prior to analysis, Brainmint RSFC data was adjusted for age, sex, tSNR and FD, same as HBN RSFC data prior to running PLS. This was done by using FSLNets’ “nets_unconfound” in MATLAB (MathWorks, 2020). As a proxy for significance, connectivity loadings were thresholded at Z-scores<|3| in visualisations, akin to the procedure in our previous work using PLS.

**Figure 2.**
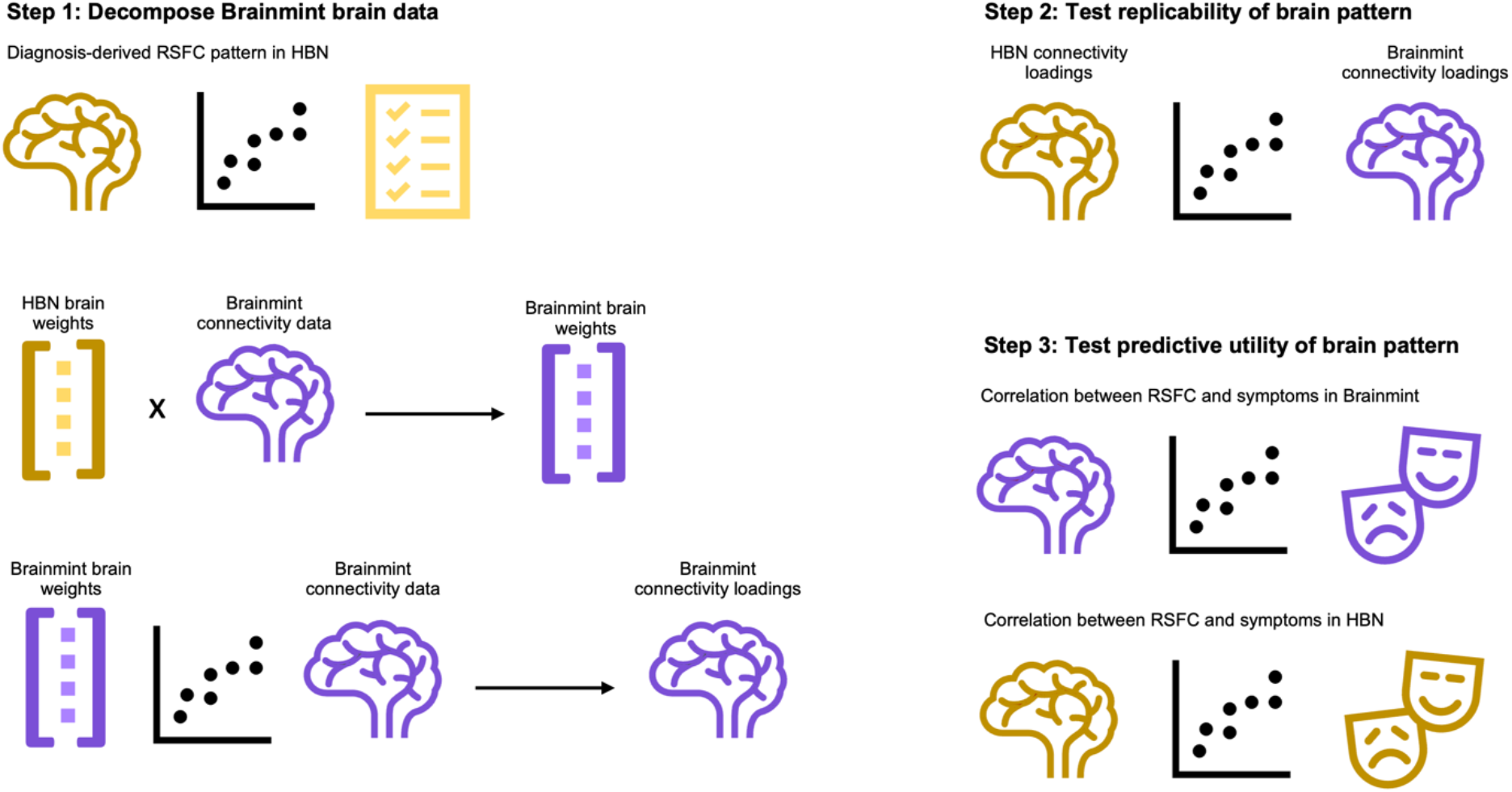
An overview of the out-of-sample validation pipeline. RSFC; resting-state functional connectivity. HBN; Healthy brain network sample. Brainmint; Brains and minds in transition sample.

#### 2.6.2 Testing the association of the brain pattern with symptom scores

To assess whether the brain pattern was associated with symptom scores, we correlated the derived Brainmint RSFC pattern with Brainmint symptom data. Specifically, we investigated the Spearman’s rank correlation between diagnosis-derived brain weights and symptom dimensions from SDQ. To assess the reliability of the associations between brain weights and symptom dimensions, we ran 1000 bootstraps using resampling with replacement. Reliability was defined as whether bootstrapped (with replacement) 95% confidence interval overlapped with zero (i.e., not overlapping with zero being considered reliable). Finally, as a test of external validity, we also ran these correlations between the HBN RSFC pattern and SDQ symptom dimensions in HBN.

## 3 Results

The correlation between Brainmint and HBN connectivity loadings was significant for both the cross-diagnostic pattern (r=.39, permuted *p*<.001; see Figure 3A) and ASD (r=.49, permuted *p*<.001; see Figure 4A), indicating that both brain patterns were validated across samples. As shown in Figure 3B, the cross-diagnostic RSFC pattern implicated weaker connectivity within the control network, in addition to weaker between-network connectivity between the salience network and control network, as well as between the default mode network (DMN) and limbic network. In terms of symptom dimensions, this connectivity pattern exhibited significant positive associations with anxiety, conduct problems, hyperactivity, and peer problems in HBN, as well as a negative association with prosocial behaviour (see Figure 3C). In Brainmint, there were no significant associations between the cross-diagnostic connectivity pattern and symptom dimensions (see Figure 3D).

**Figure 3.**
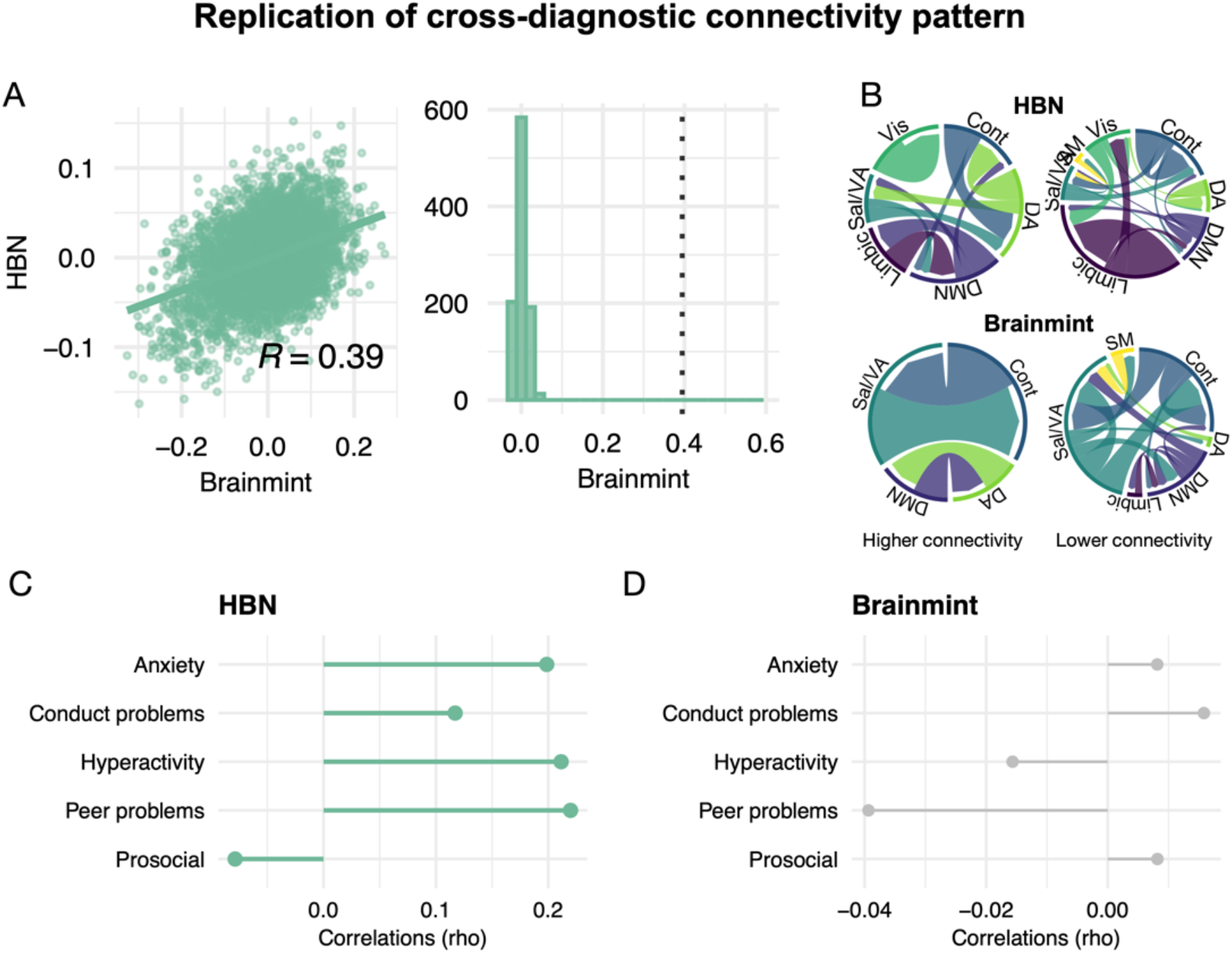
Validation of the cross-diagnostic connectivity pattern from HBN to Brainmint. **A**. Pearson’s correlation of connectivity weights between samples (left) and corresponding permutation test (right). The dotted line marks the non-permuted R value. **B**. Visualisation of RSFC pattern in each sample. Magnitude in this plot reflects summarised edge strength across each network. Depicted are thresholded edges (Z-scores<|3|). **C**. Associations between derived brain pattern and SDQ symptom dimensions in HBN. **D**. Associations between derived brain pattern and SDQ symptom dimensions in Brainmint. A positive correlation indicates higher connectivity is associated with higher symptom level and vice versa. Associations with symptom dimensions are marked in bold green if 95% confidence interval of the bootstrap distribution did not contain zero. ASD; Autism spectrum disorder. HBN; Healthy brain network sample. Brainmint; Brains and minds in transition sample. SDQ; strength and difficulties questionnaire.

**Figure 4.**
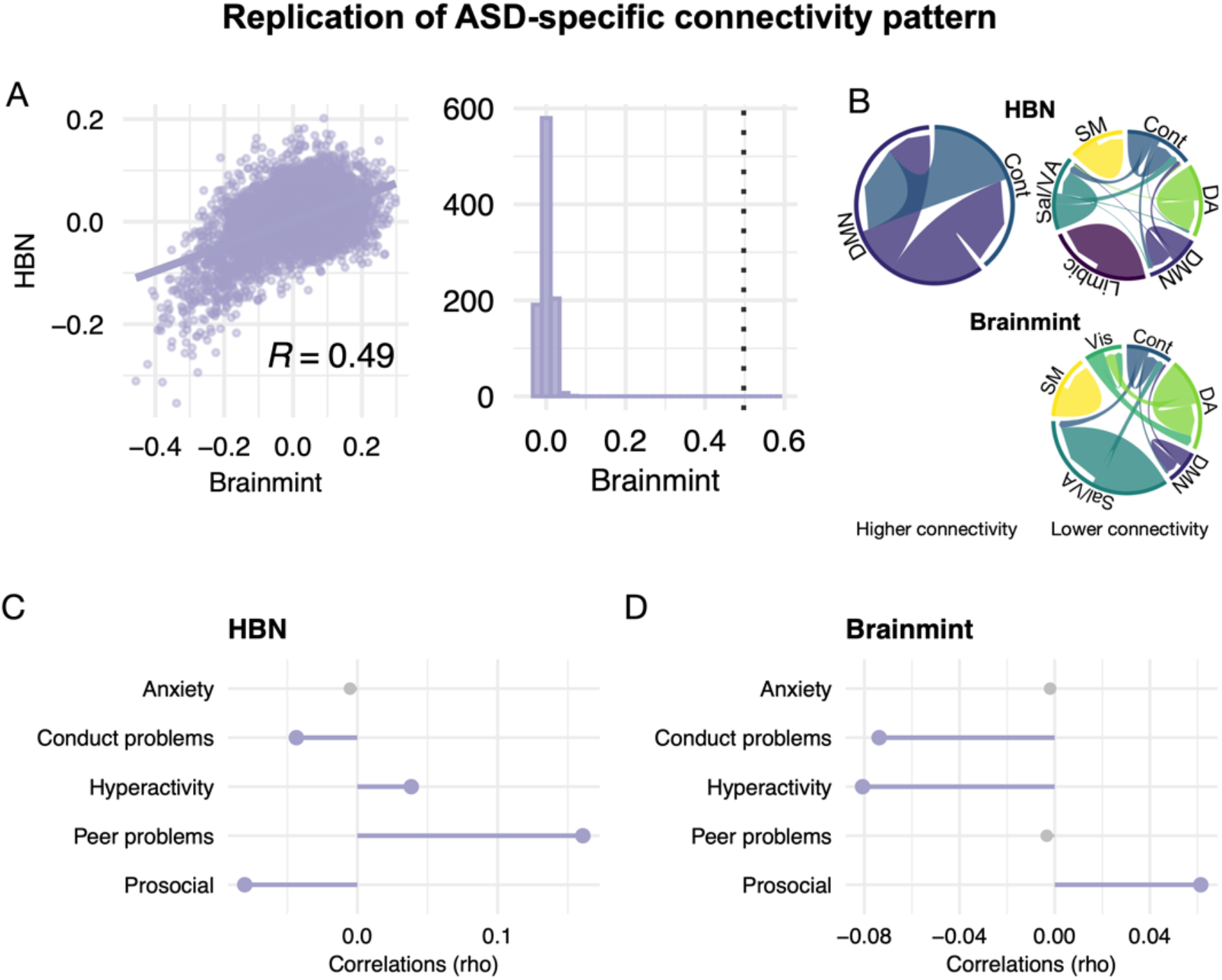
Validation of the ASD-specific connectivity pattern from HBN to Brainmint. **A**. Pearson’s correlation of connectivity weights between samples (left) and corresponding permutation test (right). The dotted line marks the non-permuted R value. **B**. Visualisation of RSFC pattern in each sample. Magnitude in this plot reflects summarised edge strength across each network. Depicted are thresholded edges (Z-scores<|3|). **C**. Associations between derived brain pattern and SDQ symptom dimensions in HBN. **D**. Associations between derived brain pattern and SDQ symptom dimensions in Brainmint. A positive correlation indicates higher connectivity is associated with higher symptom level and vice versa. Associations with symptom dimensions are marked in bold purple if 95% confidence interval of the bootstrap distribution did not contain zero. ASD; Autism spectrum disorder. HBN; Healthy brain network sample. Brainmint; Brains and minds in transition sample. SDQ; strength and difficulties questionnaire.

As shown in Figure 4B, the RSFC pattern for ASD implicated weaker within-network connectivity in the somatomotor network, dorsal attention (DA) network, salience network and DMN. In terms of symptom dimensions, this RSFC pattern was significantly associated with more symptoms of peer problems and hyperactivity in HBN, as well as lower degree of conduct problems and prosociality (see Figure 4C). In Brainmint, the ASD-specific RSFC pattern was associated with higher levels of prosocial behaviour and fewer symptoms of hyperactivity and conduct problems (see Figure 4D).

## 4 Discussion

The current study aimed to investigate whether psychopathology-related RSFC patterns are informative of vulnerability for mental illness in undiagnosed individuals by validating connectivity patterns derived in a developmental clinical sample from the US in a Norwegian convenience-based sample of youth. However, the RSFC pattern was only sensitive to symptom burden in the discovery sample. Specifically, we found that the RSFC patterns were associated with symptom dimensions thematically overlapping with core symptom characteristics in the discovery sample, while in the validation sample we found a weak, inverse relationship for ASD. This latter association was small at the group-level, with great variation at the individual level. Taken together, these results show that although diagnosis-derived RSFC patterns replicate across samples, their utility is greatly limited due to their lack of sensitivity to symptom burden.

Replicability and generalisability of neuroimaging findings remain a challenge in the field (Botvinik-Nezer & Wager, 2022). Historically, small sample sizes and lack of methodological rigour have resulted in poor replication rates, possibly reflecting that many published findings are potential false positives (Ioannidis, 2005). To overcome such challenges, increasing effort has been put into developing procedures for reproducible science (Niso et al., 2022). With the advent of multivariate machine learning approaches in neuroscience, issues related to statistical power and extensive univariate testing in small samples have been improved (Botvinik-Nezer & Wager, 2022). However, these multivariate approaches come with new challenges, such as data leakage, overfitting and the need for sufficiently large data sets to ensure robustness (Botvinik-Nezer & Wager, 2022; Davatzikos, 2019; Poldrack et al., 2020; Varoquaux, 2018). Although the current study aimed to overcome some of these challenges, we still did not obtain generalisable results.

Generalisation of RSFC patterns related to psychopathology has been hampered by challenges related to the stability and reliability of RSFC results, as well as variations in mental health profiles across cohorts (Uddin et al., 2017). Data driven approaches to symptom clustering have to some degree yielded reproducible clusters or hierarchies of symptom structure across samples (Caspi et al., 2014). Recently, we derived brain-based latent dimensions of psychopathology using symptom covariance with functional brain networks in HBN (Voldsbekk et al., 2023). Similar brain-based dimensions of psychopathology were identified in ABCD, combining measures of both brain structure and RSFC (Kebets et al., 2023). By decomposing the HBN data using feature weights estimated in ABCD from Kebets et al., we found that the symptom dimensions replicated, however the RSFC patterns did not (Voldsbekk et al., 2023). Previous attempts at replication of brain-symptom mapping in independent samples have shown similar findings of replicating latent clinical dimensions, but weak or non-replicable RSFC patterns (Linke et al., 2021). In light of this, it is surprising that the current study could validate diagnosis-based case-control and ASD-specific RSFC patterns from HBN to Brainmint. Of course, the validated RSFC pattern was not fully overlapping across the two cohorts. For example, notable differences can be observed in the higher connectivity profile associated with the cross-diagnostic pattern, in which HBN is exhibiting a greater number of networks involved compared to Brainmint. In addition, the clinical associations of this pattern in the discovery sample were not reproduced in the validation sample.

The current validation effort is conducted across two widely different samples, both in terms of age, sex distribution, and other demographical variables, and in their mental health profile. While the HBN sample consists of mainly children with diagnosed neurodevelopmental disorders, the Brainmint sample consists of adolescents recruited from the community. Some of these adolescents have elevated symptom burden and may meet the diagnostic criteria of a mental disorder, but this sample is not enriched with diagnoses as is the case with HBN. The symptom distribution is also different, with higher prevalence of anxiety symptoms in Brainmint, as compared to conduct problems and peer problems in HBN. In line with this, there is also a marked difference in the sex distribution across the two samples, with higher prevalence of males in HBN and the majority being female in Brainmint. Given these differences, it is all the more striking that the RSFC patterns were validated. Concomitantly, this may also explain the difference in clinical associations. One possible explanation could be that the RSFC patterns do reflect some vulnerability to psychopathology, only it is not sensitive enough to be associated with mental health symptoms in a widely different sample.

Prior work seem to converge on the finding that RSFC patterns relating to psychopathology are transdiagnostic or shared across disorders (Elliott et al., 2018; Karcher et al., 2021; Kebets et al., 2023; Lees et al., 2021; Linke et al., 2021; McTeague et al., 2017; McTeague et al., 2020; Sha et al., 2019; Voldsbekk et al., 2023; Xia et al., 2018). This would suggest that if anything, RSFC patterns represent possible general markers of psychopathology rather than disorder-specific markers. This has implications for their utility as biomarkers, meaning RSFC patterns may be used to detect general vulnerability, but not vulnerability specific to specific disorders. In line with this, a recent systematic review based on studies only of the general population conclude that general psychopathology is related to various RSFC patterns across studies (Hoy et al., 2023). With regards to specific psychopathology, there was only one finding reported across more than one study, namely an association between a neurodevelopmental symptom dimension and lower connectivity within the DMN (Karcher et al., 2021; Modabbernia et al., 2022). These findings are strikingly consistent with our previous work in HBN – a developmental clinical sample (Voldsbekk et al., 2023). This suggests that RSFC-psychopathology associations should extend from clinical samples to population samples, consistent with the understanding of psychopathology as a dimensional structure. Of note, this highlights the relevance of investigating the overlap and reproducibility of RSFC-psychopathology associations from clinical to population-based samples. The current study represents one attempt at this.

Consistent with the symptom load distribution in HBN across diagnostic categories, the RSFC pattern in HBN picked up associations with peer problems, hyperactivity, and prosocial behaviour for the ASD-specific pattern and all symptom dimensions for the cross-diagnostic pattern. This represents a sanity check that the diagnosis-derived RSFC pattern in HBN picks up similar associations with symptom load as the diagnosis groups they are modelled to represent. Reliability of mental health measures has remained a challenge in the field (Nikolaidis et al., 2022). Here we show that RSFC patterns derived from diagnostic information exhibit associations with an independent measurement of symptom load that are overlapping with symptom load associations observed for each diagnostic category. This supports that these RSFC patterns reflect something that overlaps with their corresponding diagnostic categories. However, this sensitivity of the RSFC pattern was not generalisable to Brainmint. The RSFC pattern in Brainmint picked up an inverse relationship with prosocial behaviour, hyperactivity, and conduct problems for the ASD-specific pattern, indicating that higher prosociality and lower conduct problems and hyperactivity was associated with a more “ASD-like” brain pattern. This finding is paradoxical and the opposite of what one would expect. While it is too early to conclude based on one preliminary association only, it is worth noting that the strength of this association was low. Similarly, there was no significant associations with the cross-diagnostic pattern in Brainmint. Given these weak group-level associations, with great variation at the individual level, adding too much emphasis to this preliminary finding is unwarranted. Instead, this result adheres to the previous literature finding generalisation of RSFC results a challenge (Uddin et al., 2017).

Some further limitations should be noted. First, RSFC results are influenced by methodological choices (Sala-Llonch et al., 2019; Shirer et al., 2015). To increase reproducibility of RSFC networks, we utilised an established parcellation scheme (Schaefer et al., 2018). Second, the two samples underwent slightly differing fMRI preprocessing pipelines. This difference should diminish, rather than inflate, any ability to reproduce findings, meaning that the current study represents a conservative approach to reproducibility with higher chance of false negatives than false positives. Third, the symptom dimensions from SDQ were measured by parent-report in HBN and by self-report in Brainmint. This difference may have induced systematic variations in the data across the two samples, as it is known that parent-report and self-report have small-to-moderate correlations (Gaete et al., 2018). This limitation represents a possible explanation for why we did not find overlapping associations between RSFC patterns and symptom level across the two cohorts. In addition, this may explain why the community-based sample seemed to exhibit a higher symptom burden than the clinical sample. This limitation represents an important reminder that low reliability in mental health measures impedes scientific discovery (Nikolaidis et al., 2022) and underscores that the current results must be interpreted with caution. Fourth, the age, sex, and clinical distributions across the two samples differed, meaning we cannot rule out whether the lack of generalisability across samples reflect these differences in distributions rather than a lack of reproducibility of RSFC-symptom patterns per se.

## 5 Conclusions

This work demonstrates that diagnosis-derived RSFC patterns in a US developmental clinical sample can be extended to a Norwegian convenience-based sample of youth. Both the cross-diagnostic and the ASD-specific RSFC patterns were validated across samples. However, although both connectivity patterns exhibited significant associations with thematically appropriate symptom dimensions in both the discovery sample (HBN), they were not found to be sensitive to overlapping dimensions of symptom burden in the validation sample (Brainmint). Implications of this work is that generalisation of RSFC results remains a challenge. For any psychopathology-related RSFC patterns to be generalisable and clinically relevant, their sensitivity to symptom burden across samples represents a prerequisite.

## Data Availability

The code and data used in the study will be made available upon publication in a public repository (Open Science Framework) (https://osf.io/cdjw4/).

## Acknowledgements

This project was funded by research grants from the Research Council of Norway (Grant Nos. L.T.W: 249795, 273345, 300767), the South-Eastern Norway Regional Health Authority (Grant Nos. L.T.W: 2014097, 2015073, 2016083, 2018076, 2019101. D.A: 2019107, 2020086), KG Jebsen Stiftelsen, ERA-Net Cofund through the ERA PerMedproject IMPLEMENT, the Horizon Europe HORIZON-HLTH-2021-STAYHLTH-01 (101057429), and the European Research Council under the European Union s Horizon 2020 research and Innovation program (L.T.W: ERC StG Grant No. 802998).

The work was performed on the Service for Sensitive Data (TSD) platform, owned by the University of Oslo, operated, and developed by the TSD service group at the University of Oslo IT-Department (USIT). Computations were also performed using resources provided by UNINETT Sigma2—the National Infrastructure for High Performance Computing and Data Storage in Norway.

## Notes

### Competing Interest Statement

The authors have declared no competing interest.

### Author Declarations

We hereby confirm that Regional Committee for Medical & Health Research Ethics, Section A, South East Norway, approved the Research Project Brains and minds in transition (BRAINMINT): Parsing the heterogeneous developmental and genetic architecture of risk and resilience in the adolescent brain on the 18th of December 2019. The Project Manager for the study is Lars Tjelta Westlye and the Institution Responsible for Research is the University of Oslo. The approval has been given on the basis that Research Project will be implemented as described in the Research Protocol.

### Summary of Updates

Clarification

