## Supplementary material for "Testing the sensitivity of diagnosis-derived patterns in functional brain networks to symptom burden in a Norwegian youth sample"

### Content

- Supplementary Figure 1. SDQ symptom distributions across diagnostic categories in HBN
- Supplementary Table 1. Demographic table with clinical values scoring over cut-off
- Supplementary references

**Supplementary Figure 1. SDQ symptom distributions across diagnostic categories in HBN**

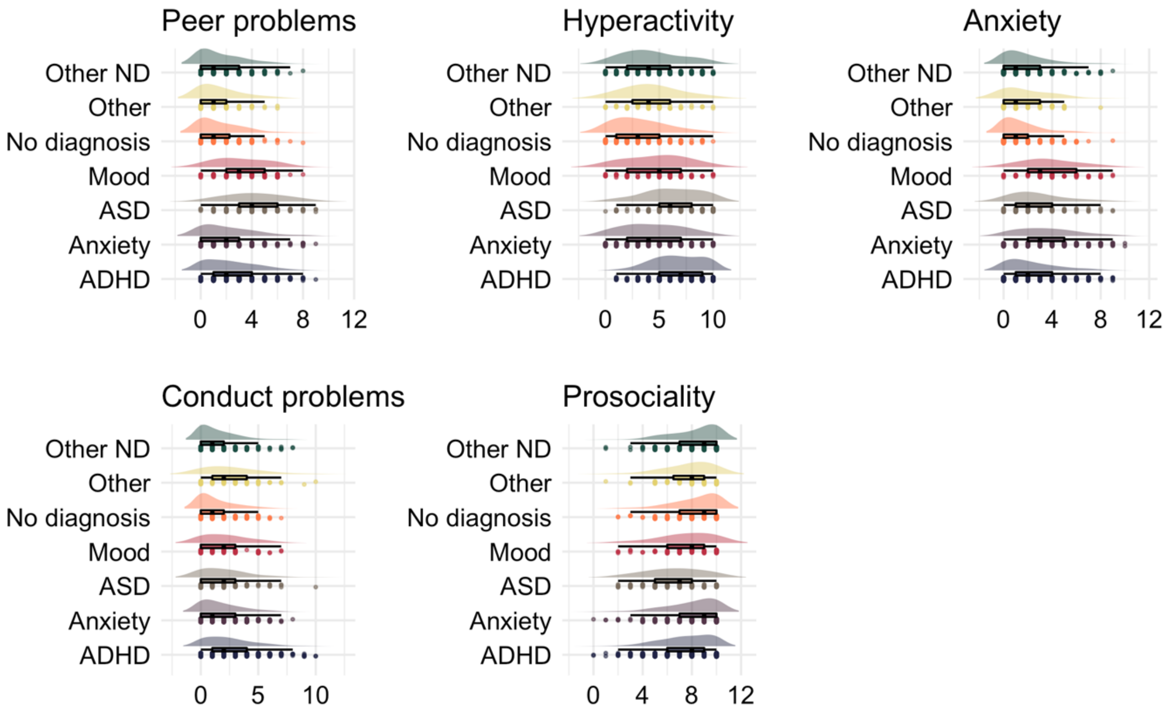

**Supplementary Table 1. Demographic table with clinical values scoring over cut-off**

| | HBN (n = 1666) | Brainmint (n = 531) |
|---|---|---|
| **Demographic information** | | |
| Age | 10.91 ± 3.14 | 17.69 ± 2.83 |
| Sex (n female) | 641 | 390 |
| **SDQ sumscores** | | |
| Peer problems | 2.19 ± 2.06 [419] | 2.54 ± 1.88 [136] |
| Hyperactivity | 5.3 ± 2.84 [599] | 4.38 ± 2.58 [121] |
| Anxiety | 2.43 ± 2.23 [301] | 4.4 ± 2.68 [245] |
| Conduct problems | 1.96 ± 1.96 [321] | 1.61 ± 1.49 [59] |
| Prosociality | 7.78 ± 2.09 [124] | 8.56 ± 1.56 [10] |

*Note*. Mean ± standard deviation [Number of participants scoring above clinical cut-off] (Bryant et al., 2020). HBN; Healthy brain network. Brainmint; Brains and minds in transition study. SDQ; Strength and difficulties questionnaire.
